# Implementation of an in-house real-time reverse transcription-PCR assay to detect the emerging SARS-CoV-2 N501Y variants

**DOI:** 10.1101/2021.02.03.21250661

**Authors:** Marielle Bedotto, Pierre-Edouard Fournier, Linda Houhamdi, Philippe Colson, Didier Raoult

## Abstract

The SARS-CoV-2 pandemic has been associated with the emergence of several variants with a mutated spike glycoprotein that are of substantial concern regarding their transmissibility and ability to evade immune responses. This warrants implementing strategies for their detection and surveillance. We have set up an in-house one-step real-time reverse transcription-PCR (qPCR) assay that specifically detects SARS-CoV-2 spike N501Y variants. Our assay was positive for all 6 patients found spike N501Y-positive by genome sequencing. Ten cDNA samples for each of the 10 Marseille variants identified by genome sequencing and three nasopharyngeal samples of a spike N501Y-negative variant (Marseille-4) that predominates locally tested negative. All negative controls among which 5 SARS-CoV-2-negative nasopharyngeal samples tested negative. First use in the setting of diagnosis on 51 nasopharyngeal samples from SARS-CoV-2-positive but Marseille-4-negative patients showed positivity in 5 cases further confirmed by sequencing as from spike N501Y variant-infected patients. Thus, our in-house qPCR system was found reliable for the detection of the N501Y substitution and allowed estimating preliminarily that spike N501Y variant prevalence was 4% among SARS-CoV-2 diagnoses since January 2020.

## TEXT

The SARS-CoV-2 pandemic has been associated with the occurrence of several viral variants with a mutated spike glycoprotein (S). Those currently of greatest concern carry N501Y substitution within the spike (S) receptor binding domain (RBD). Indeed, they have become predominant in England (20I/501Y.V1) [1] and were detected in South Africa (20H/501Y.V2) [2] and Brazil [3]. The 20I/501Y.V1 variant has started to spread worldwide including in France [4]. It has been reported as 50-74% more transmissible than preexisting strains, suspected to evade anti-spike antibodies [1], and it caused a reinfection [5]. Its real-time detection is critical to manage patients appropriately, monitor and assess its epidemiological and clinical features, and survey cases of immune escape post-infection or vaccination. An alternative comprehensive detection strategy is warranted considering the very large number of SARS-CoV-2 cases.

We have implemented an in-house one-step real-time reverse transcription-PCR (qPCR) assay that specifically detects SARS-CoV-2 N501Y variants by targeting nucleotide position 23,063 within S gene where A>U leads to N501Y. SARS-CoV-2 genomes from the GISAID database with or without N501Y were used to design primers and a hydrolysis probe. The sequences of these latter and the qPCR conditions are shown in Table 1. Our N501Y-specific assay was positive for all 6 patients identified as 20I/501Y.V1-positive by genome sequencing and for all 3 additional contact patients for whom sequences were not obtained due to low viral load (Ct of qPCR diagnosis test= 32.0-34.0) leading to no amplification by two conventional PCR. Ten cDNA samples, one for each of the 10 Marseille variants [6] identified by genome sequencing, and three additional nasopharyngeal (NP) samples of the Marseille-4 variant that predominates locally (≈two-thirds of diagnoses) tested negative. All negative controls (5 SARS-CoV-2-negative NP samples and 20 RNA extract-free qPCR mixes) tested negative. The first use for diagnosis purpose conducted on 51 NP samples from SARS-CoV-2 positive (Ct= 10.2-30.8) but Marseille-4-negative patients showed positivity for 5 samples, which were confirmed by sequencing as from N501Y variant-infected patients.

**Table 1.**
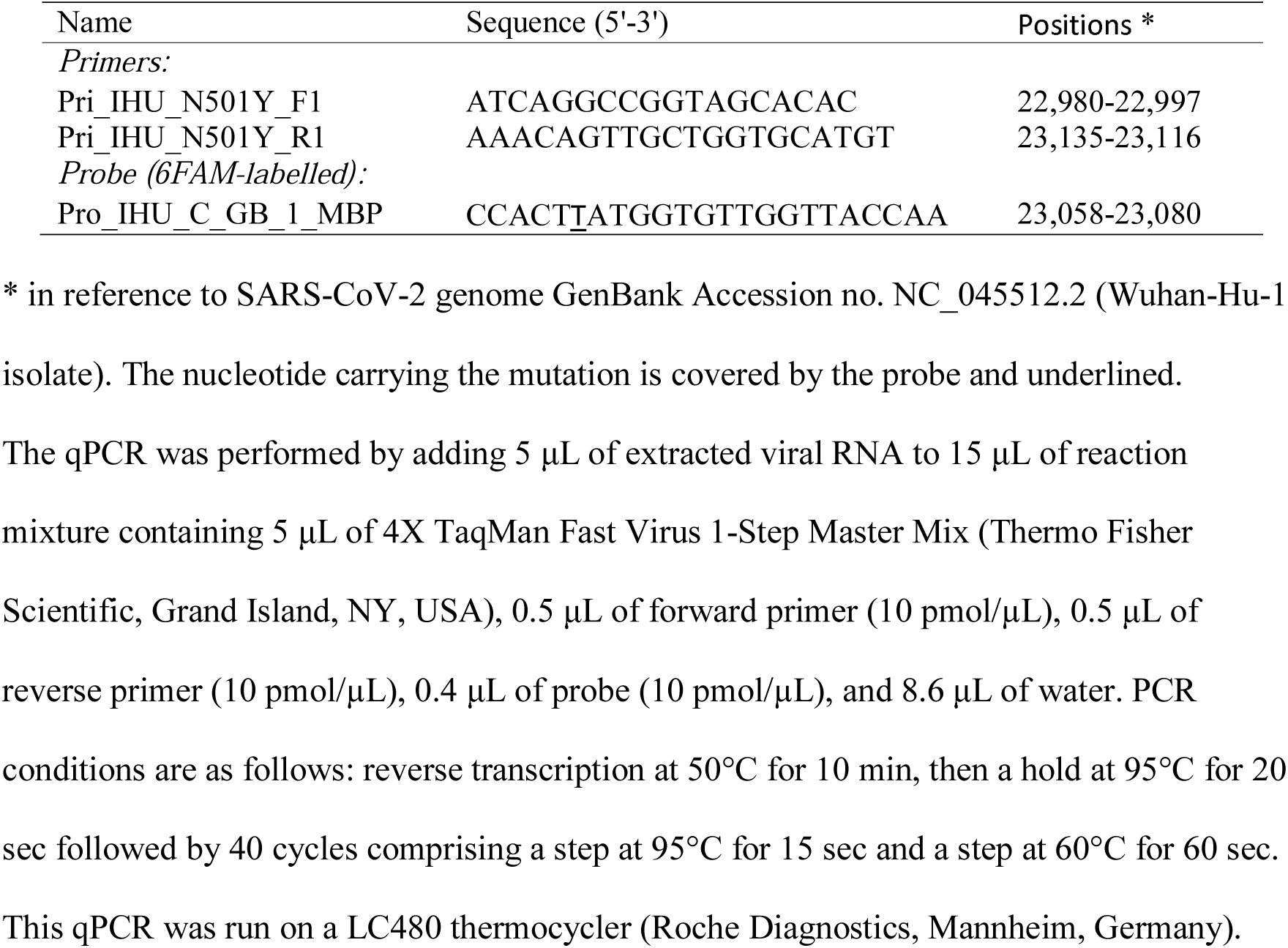
Primers, probe and qPCR conditions

Our in-house qPCR system was found reliable to detect specifically the N501Y substitution and preliminarily allowed estimating 20I/501Y.V1 variant prevalence to 4% among our current SARS-CoV-2 diagnoses since January. This assay showed better sensitivity than conventional PCR and should be able to detect different N501Y variants. A commercialized RT-PCR diagnosis assay (TaqPath RT-QPCR test) allows the indirect identification of the 20I/501Y.V1 variant by detecting its ORF1a and N genes but not its S gene due to a deletion at positions 21,766-21,772 [4]. However this deletion is also present in strains devoid of the N501Y substitution and was reported in 0.6% of recent SARS-CoV-2 diagnoses in France [4]. Moreover, it is absent from the South African and Brazilian N501Y variants, which prevents their identification. Finally, our in-house qPCR test can be widely and easily deployed in laboratories as it runs on any open qPCR microplate platform, does not require technical workers’ training, and is as cheap as other in-house qPCR assays. Such approach should allow adapting continuously diagnosis strategies for new SARS-CoV-2 variants.

## Data Availability

Data underlying the study are available from the corresponding author upon request.

## Author contributions

Conceived and designed the study: PC, DR. Contributed materials/analysis tools: MB, PEF, LH, PC. Analyzed the data: all authors. Wrote the paper: PC, DR.

## Acknowledgments -Funding

This work was supported by the French Government under the “Investments for the Future” program managed by the National Agency for Research (ANR), Méditerranée-Infection 10-IAHU-03 and was also supported by Région Provence Alpes Côte d’Azur and European funding FEDER PRIMMI (Fonds Européen de Développement Régional-Plateformes de Recherche et d’Innovation Mutualisées Méditerranée Infection), FEDER PA 0000320 PRIMMI.

## Conflicts of interest

The authors declare that they have no known competing financial interests or personal relationships that could have appeared to influence the work reported in this paper. Funding sources had no role in the design and conduct of the study; collection, management, analysis, and interpretation of the data; and preparation, review, or approval of the manuscript.

## Ethics

This study has been approved by the ethics committee of the University Hospital Institute Méditerranée Infection, Marseille, France, with the registration number 2020-029.

## Notes

### Competing Interest Statement

The authors have declared no competing interest.

### Author Declarations

This study has been approved by the ethics committee of the University Hospital Institute Mediterranee Infection, Marseille, France, with the registration number 2020-029.

